# icognition: a smartphone-based cognitive screening battery

**DOI:** 10.1101/2023.07.19.23292824

**Authors:** Stijn Denissen, Delphine Van Laethem, Johan Baijot, Lars Costers, Annabel Descamps, Ann Van Remoortel, Annick Van Merhaegen-Wieleman, Marie B D’hooghe, Miguel D’Haeseleer, Dirk Smeets, Diana Maria Sima, Jeroen Van Schependom, Guy Nagels

## Abstract

**Background:** Telemedicine is feasible and well-accepted by people with multiple sclerosis (MS).

**Objective:** The aim of this study is to validate a smartphone-based cognitive screening battery, ico**gnition**, to faster signal cognitive deterioration.

**Methods:** ico**gnition** consists of three tests (Symbol Test, Dot Test and visual Backwards Digit Span (vBDS)) that are equivalents of validated paper-pencil tests. These are the Symbol Digit Modalities Test (SDMT), the 10/36 Spatial Recall Test (SPART) and the auditory Backwards Digit Span (aBDS), respectively. To establish the validity of ico**gnition**, 101 people with MS and 82 healthy subjects completed all tests. 21 healthy subjects repeated testing 2 to 3 weeks later.

**Results:** All tests in ico**gnition** correlate well with their paper-pencil equivalent (Symbol Test: r=.63, p<.001; Dot Test: r=.31, p=0.002; vBDS: r=.71, p<.001), negatively correlate with the Expanded Disability Status Scale (EDSS: Symbol Test: rho=-.27, p=.01; Dot Test: rho=-.29, p=.006; vBDS: rho=- .23, p=.027) and show high test-retest reliability (Symbol Test: r=.81, p<.001; Dot Test: r=.75, p<.001; vBDS: r=.84, p<.001). Test performance was not significantly different between people with MS and healthy subjects for all cognitive tests, both in ico**gnition** and their paper-pencil equivalents.

**Conclusion:** ico**gnition** is a valid and reliable tool to remotely screen for cognitive functioning in persons with MS.

## Introduction

Medicine is increasingly digitalising, and there are solid reasons to stimulate this trend. Doctors can more easily access and share electronic health records, and storing data in a digital format facilitates visualisation and organisation in research databases, yielding new insights into pathology and disease management. Beyond a nice-to-have, digital medicine was a crucial element in handling the COVID-19 pandemic, allowing telemedicine to be practiced when social distancing was necessary.

Telemedicine provides practical solutions for people with multiple sclerosis (MS). Telemedicine tools are well-accepted by patients^1,2^, and their feasibility^2^ and cost-effectiveness^1^ have been established previously. Patients moreover tend to objectively benefit from these approaches, for example for fatigue management^3^ and improving cognitive function^4^. The latter is important as nearly half of the people with MS have cognitive impairment^5^, which is associated with significant financial burden^6^, productivity^7^, and decreased mental well-being^8^. It is hypothesised that cognitive problems in multiple sclerosis start with slowed information processing speed^9^. It is therefore promising that digital solutions for assessing this domain are currently emerging^10,11^.

Existing digital cognitive tests for people with MS however have two important limitations; they are mainly created for tablets^10,11^ and do not capture memory impairment, which is even more prevalent^12^. Compared to smartphones, tablets offer the advantage of a bigger screen size, at the cost of people using them far less frequently. Some cognitive tests were designed for smartphones, such as the cognitive test in Floodlight^13^ and the one presented by Pham et al. 2021^14^. These tests are a digital version of the symbol digit modalities test (SDMT)^15^, a fair choice considering its ease of use and excellent psychometric properties^16^. Digitalising the SDMT allows randomising its key, which could reduce practice effect as reported in Pereira et al. 2015^17^. Creating an exact digital replica of the SDMT however requires patients to choose from nine tiny buttons on the screen, which could cause motor interference as fine motor skills are commonly affected in MS^18^.

This study aims to validate a new smartphone-based screening battery called “ico**gnition**”, for monitoring cognitive problems in people with MS. The application is intended for remote follow-up of cognitive performance by regularly screening for cognitive deterioration. It is intended to be part of the recently established ico**mpanion** application^19^. Regular remote screening could lead to a more prompt reaction in terms of confirming deterioration by a neuropsychologist and addressing this deterioration by the patient’s neurologist.

## Methods

### Participants

Inclusion criteria for people with MS were a confirmed diagnosis of multiple sclerosis according to the McDonald criteria^20^. MS subjects were excluded if they were hospitalised, with the exception of the underlying reason being rehabilitation. Both persons with MS and healthy control subjects were excluded in the presence of any other neurological or psychiatric disorder or learning disorder. A total of 101 people with MS and 82 healthy control subjects met the in- and exclusion criteria for this study. All subjects were either Dutch-speaking or bilingual including Dutch and were 18 years or older.

### Ethics

This study was approved by the “Commissie Medische Ethiek” of the UZ Brussel (B.U.N. 143201940335) and the National MS Center of Melsbroek. All participants signed informed consent prior to inclusion.

### icognition

The ico**gnition** cognitive screening battery consists of three tests.

The Symbol Test (figure 1) is based on the computerised digit-symbol substitution test (DSST) presented in Rypma et al. 2006^21^. A combination of symbols is presented to the subject, one at a time. Furthermore, a key is presented on top, which consists of 9 pairs of symbols, and which shuffles every trial. For every trial, the subject needs to indicate whether the combination occurs in the key on top. The total score is the number of correct answers in 90 seconds. This test is designed to capture information processing speed.

**Figure 1:**
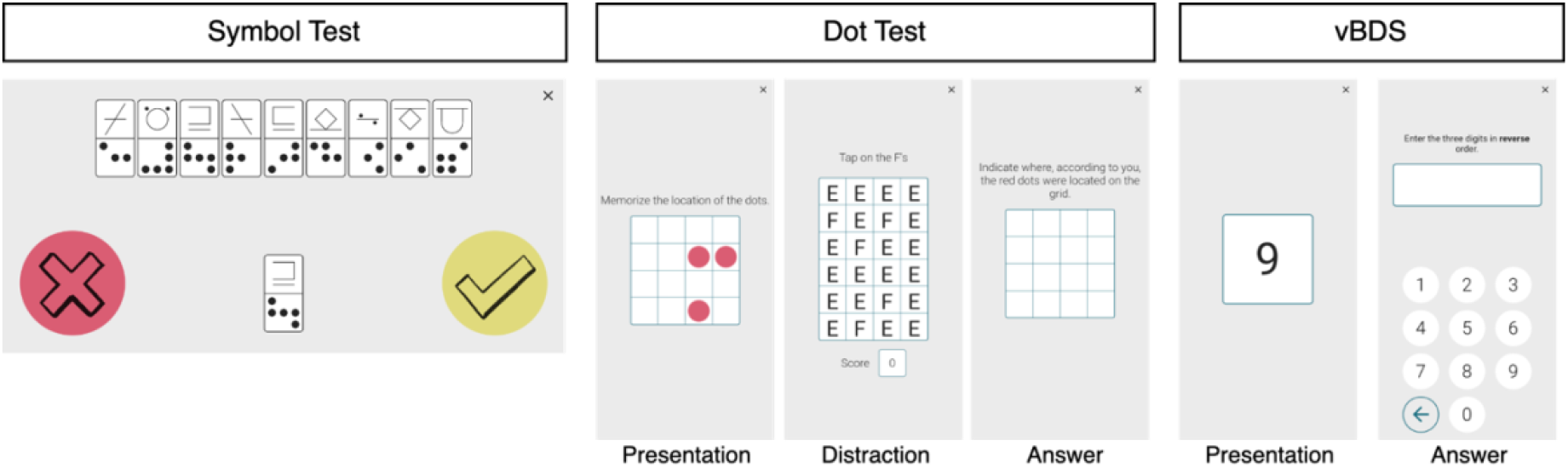
Screenshots of the ico**gnition** tests. Note: although the instructions were in Dutch during testing, they are presented here in English. vBDS = visual Backwards Digit Span.

The Dot Test (figure 1) consists of three phases. In a first phase, a subject is presented a 4×4 grid in which three dots are shown for 3 seconds. Then, as a means of distraction, the subject is presented a 4×6 grid of “E” and “F” shapes and has to identify as many “F” shapes as possible in 4 seconds. In the last phase, the subject has to indicate where the three dots of the first phase were located in an empty 4×4 grid. The Dot Test is inspired by the Dot Memory Test presented in Sliwinski et al. 2016^22^. More specifically, all grids were reduced in size compared to their version (5×5). We also implemented a criterion for the distractor task, namely that at least 3 F shapes are identified. If this criterion is not met, the trial is restarted. The three dots could not be on one line or in an L-shape within a 2×2 block of cells. The total score is the number of correctly indicated dots across 10 trials. This test is designed to capture visuospatial short-term memory and learning.

The visual Backwards Digit Span (vBDS) test (figure 1) was inspired by the backwards digit span of the Wechsler Adult Intelligence Scale IV (WAIS-IV)^23^. As discussed in Woods et al. 2011^24^, in the original design (discussed later in the methods section), subjects with the same score can have a different number of correct spans. To mitigate this, we used a fixed number of spans. Spans were randomly generated with digits between 0 and 9, using the restraints that a digit can only occur once in the span and that a chain of three or more digits could not have a fixed increment or decrement of 1 or 2, according to Woods et al. 2011^25^. A series of digits appears on the screen one by one for 1 second per digit, consistent with Hilbert et al. 2015^26^. The subject then needs to list the digits in reversed order. Scoring consists of calculating the sum of all correct span lengths. For example, if a subject enters two spans of 3 and 1 of 4 correctly, the total score is 10. This test is designed to capture working memory.

All tests were performed on a Samsung Galaxy A10 (6.2 inch screen size). The Symbol Test needs to be performed in landscape position of the smartphone, whereas for the Dot Test and Backwards Digit Span, the smartphone needs to be in portrait position.

### The validation procedure

We used the 5-step validation process described in Benedict et al. 2012^27^ to validate ico**gnition**. Each step is addressed below.

Step 1 and 2: Standardization. In this light, the authors mention that tests should have consistent stimulus presentation and good face validity. The consistency of stimulus presentation was assured by design of the application, with prespecified (cfr. supra) time blocks for each phase of a test.

Although there is no formal criterion for face validity (the extent to which a test measures what it appears to measure), we established the concurrent validity of each ico**gnition** test by assessing how well they correlate with their paper-pencil equivalent. For the Symbol Test, this was the symbol-digit modalities test (SDMT)^15^. A sheet is presented to the subject with a key of 9 symbol-digit pairs on top and a list of symbols without a digit. In 90 seconds, the subject needs to convert as many symbols to digits as possible from the list, reading them out loud to the examiner, using the key on top. The Dot Test is based on the 10/36 spatial recall test (SPART)^28^. Here, the subject is presented a 6×6 grid with a pattern of 10 dots. This pattern is presented for 10 seconds. Subsequently, the grid is removed, and the subject is asked to replicate the pattern using 10 checkers. This cycle is repeated three times. The final score is the total number of correctly placed checkers across all trials. Finally, for the Backwards Digit Span, a modified version of the WAIS-IV auditory backwards digit span test^23^ was used. We will refer to this test as “auditory Backwards Digit Span (aBDS))”. A fixed list of spans was used, ranging in length from 3 to 7 (table S2); the complete list was always administered for each subject. The scoring metric is the same as described earlier in the ico**gnition** Backwards Digit Span.

As all subjects were Dutch-speaking, test instructions were in Dutch, and no further interference of language is expected since all tests rely on visual stimuli. Further measures to assure standardisation were maximising screen brightness and audio volume, minimising distractors by enabling “airplane mode” and “Do Not Disturb” mode, and reducing other distracting stimuli in the testing environment. Lastly, to assure the correct position of the smartphone for each test, instructions were implemented to rotate the phone in case of incorrect positioning.

Step 3: Normalisation. We tested 82 healthy controls matched on age, sex, and education level.

Step 4: Test-retest reliability. The authors mention that this criterion should be assessed on a “small sample” of either MS patients or healthy controls. We targeted to retest HC subjects 2-3 weeks after baseline testing.

Step 5: Criterion validity. For each test, we compared the performance of MS subjects to those of age-, sex- and education level-matched healthy control subjects. For this, we used a Mann-Whitney U test. We also established criterion validity by using regression-based norms, for which the analysis and results can be consulted in the supplementary material.

In addition, ecological validity was assessed with the correlation between each ico**gnition** test and the Expanded Disability Status Scale (EDSS)^29^, disease duration, the Beck Depression Inventory (BDI)^30^ and the Fatigue Scale for Motor and Cognitive Functions (FSMC)^31^.

### Data curation

Data were entered independently by two researchers (SD and DVL). Conflicts in data entry were resolved through mutual discussion.

## Statistical analyses

We used an alpha level of 0.05 for all analyses throughout this paper. Subjects having missing data on a certain test were only excluded for that specific test. We used Spearman correlation for non-linear scales (EDSS, BDI, and FSMC) for ecological validity, whereas Pearson correlation was used for disease duration. For concurrent validity and test-retest reliability, Pearson correlation was used. We used a Mann-Whitney U test for criterion validity for all ico**gnition** tests.

## Results

Both the MS and HC sample are described in Table 1. For the dot test, the first 3 subjects with MS were excluded as they were tested with an earlier ico**gnition** version where half of the trials were with three dots in a 5×5 grid. This was changed afterwards as we deemed this test to be too difficult.

**Table 1:** Group characteristics. Abbreviations: N = sample size, SD = standard deviation, IQR = Interquartile range, RRMS = Relapsing-Remitting MS, SPMS = Secondary Progressive MS, PPMS = Primary Progressive MS, EDSS = Expanded Disability Status Scale, SDMT = Symbol-Digit Modalities Test, SPART = Spatial Recall Test, BDS = Backwards Digit Span, BDI = Beck Depression Index, FSMC = Fatigue Scale for Motor and Cognitive Functions, vBDS = visual Backwards Digit Span. * Mann-Whitney U test, ** Chi-squared test. The following variables had missing values: EDSS (7), SPART (HC: 1, MS: 1), BDI (MS: 2, HC: 1), FSMC (MS: 2, HC: 7), Dot Test (MS: 3, HC: 1), vBDS (MS: 1). An additional 3 subjects were excluded for the Dot Test since they were tested on larger grid sizes.

|  | People with MS | Healthy Controls | p-value |
| --- | --- | --- | --- |
| *Demographics* |  |  |  |
| N | 101 | 82 | / |
| Age (Mean (SD)) | 45.4 (10.0) | 46.8 (14.7) | .588* |
| Sex (female:male) | 63:38 | 54:28 | .740** |
| Education (Median; IQR) | 15; 5 | 15; 4 | .739* |
| *MS-specific* |  |  |  |
| Disease duration in years (Mean (SD)) | 11.7 (74) | / | / |
| Type MS (RRMS:SPMS:PPMS) | 86:8:7 | / | / |
| EDSS (Median; IQR) | 3; 2 | / | / |
| *Paper-pencil tests* |  |  |  |
| SDMT (Mean (SD)) | 58.5 (10.0) | 58.3 (9.9) | .822* |
| 10/36 SPART (Mean (SD)) | 20.6 (4.3) | 20.1 (4.6) | .741* |
| Auditory BDS (Mean (SD)) | 48.4 (18.7) | 46.0 (17.6) | .374* |
| BDI (Mean (SD)) | 9.3 (5.7) | 5.8 (5.1) | < .001* |
| FSMC (Mean (SD)) | 58.7 (16.4) | 39.9 (12.1) | < .001* |
| *icognition tests* |  |  |  |
| Symbol Test (Mean (SD)) | 24.8 (6.3) | 25.4 (6.4) | .416 |
| Dot Test (Mean (SD)) | 21.8 (5.1) | 21.2 (4.9) | .325 |
| vBDS (Mean (SD)) | 46.9 (16.9) | 43.8 (16.7) | .268 |

### Test-retest reliability

In total, 20 HC subjects were retested with an average intertest interval of 18 days (range: 14 – 23 days) after initial testing to establish test-retest reliability. Each test in ico**gnition** showed good test-retest reliability (figure 2). Symbol Test: r = .81, p < .001; Dot Test: r = .75, p < .001; vBDS: r = .84, p < .001.

**Figure 2:**
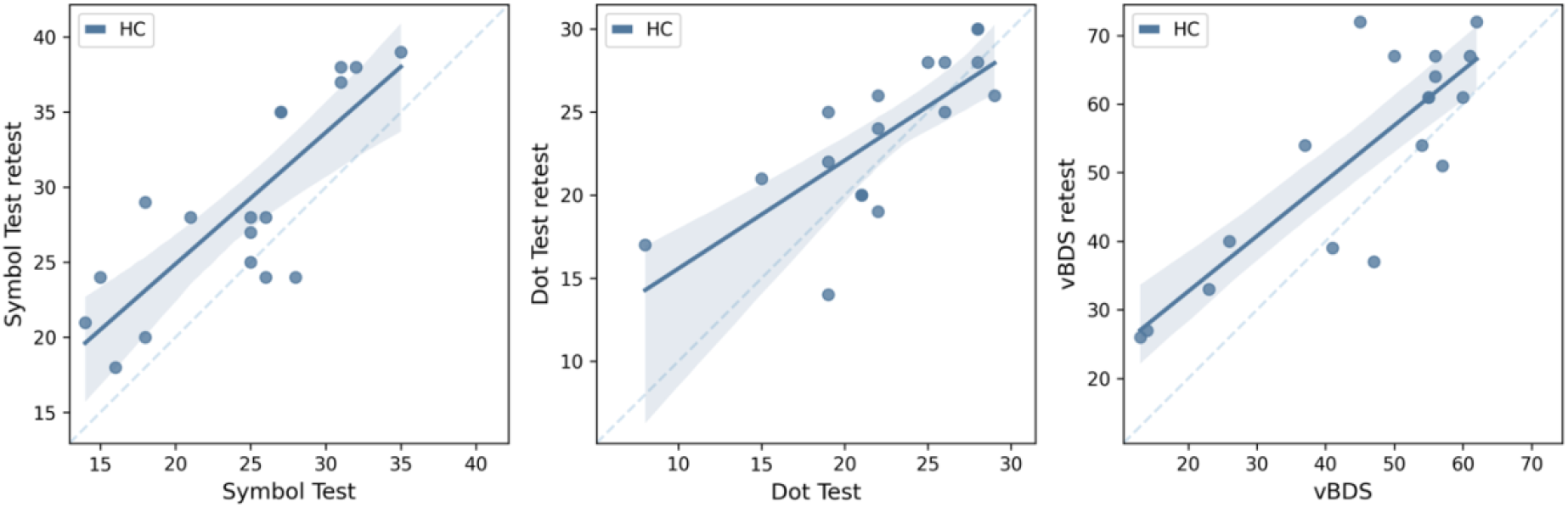
Test-retest reliability.

### Criterion validity

For all tests in ico**gnition**, there was no significant difference in performance between healthy subjects and people with MS (figure 3). Symbol Test: U = 4115.5, p = .568; Dot Test: U = 3459.5, p = .307; vBDS: U = 3506.5, p = .227.

**Figure 3:**
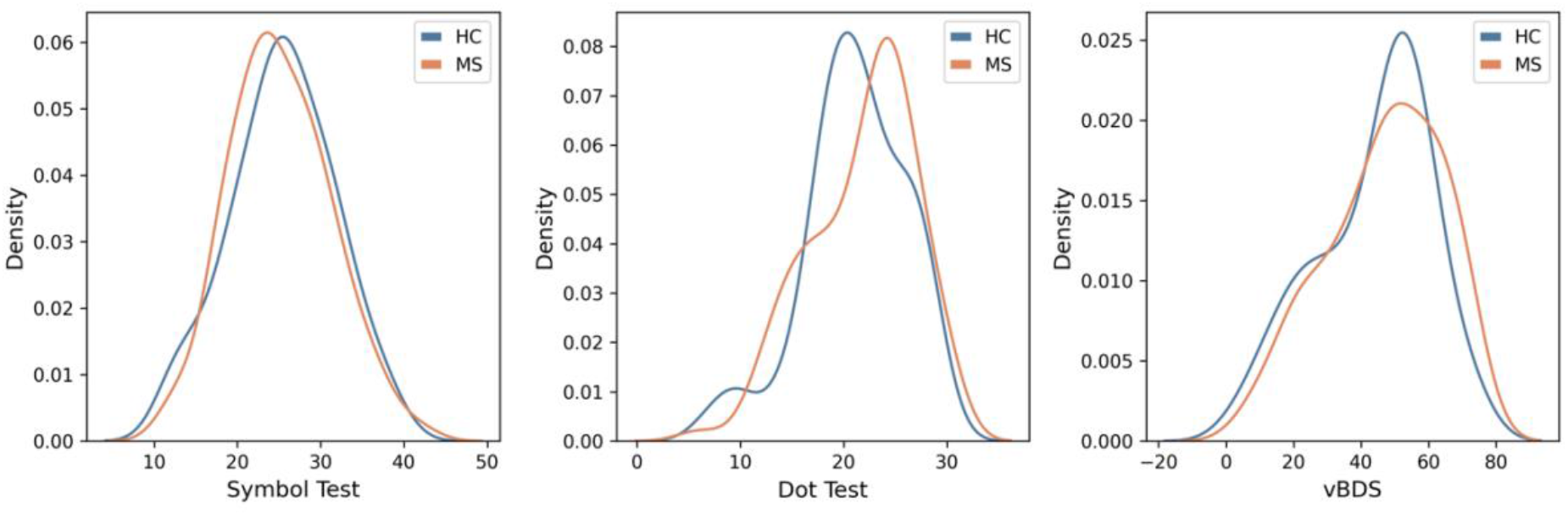
Criterion validity. Comparison of the performance of healthy subjects and people with MS on the ico**gnition** tests.

### Concurrent validity

Figure 4 shows the scatterplot of each ico**gnition** test with its paper-pencil equivalent. The Symbol Test significantly correlated with SDMT performance (HC: r = .67, p < .001; MS: r = .63, p < .001). There was also a significant correlation between the Dot Test and the SPART (HC: r = .30, p = .007; MS: r = .31, p = .002) and between the vBDS and its auditory equivalent (HC: r = .69, p < .001; MS: r = .71, p < .001).

**Figure 4:**
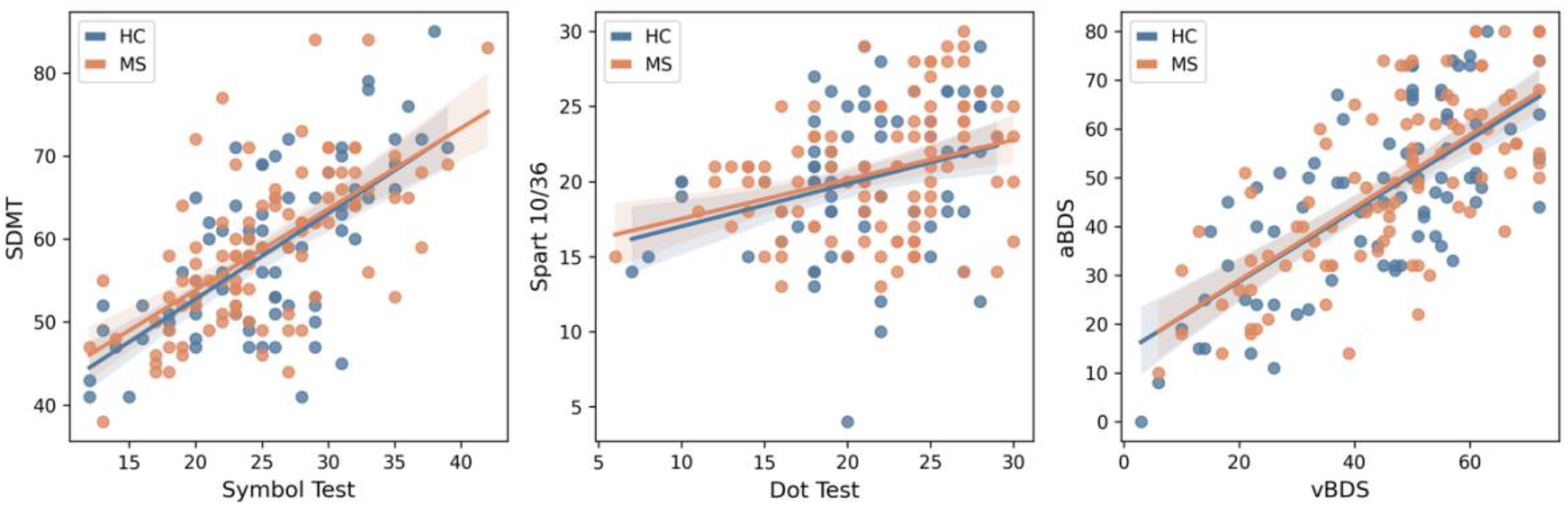
Concurrent validity. Scatterplot of each ico**gnition** test and its paper-pencil equivalent.

### Ecological validity

Patients with MS with higher EDSS scores scored worse on each ico**gnition** test (figure 5). Symbol Test: rho = -.33, p = .002; Dot Test: rho = -.32, p = .003; vBDS: rho = -.21, p = .048. Furthermore, we tested the ecological validity between each ico**gnition** test and disease duration, BDI and FSMC. All but the Symbol Test correlated with disease duration: Symbol Test: r = -.19, p = .055; Dot Test: r = - .28, p = .006; vBDS: r = -.31, p = .002. Lastly, only the Dot Test showed a significant negative relationship with the FSMC total score (rho = -.26, p = .012). For a visual representation, we refer to supplementary figure S3-S5.

**Figure 5:**
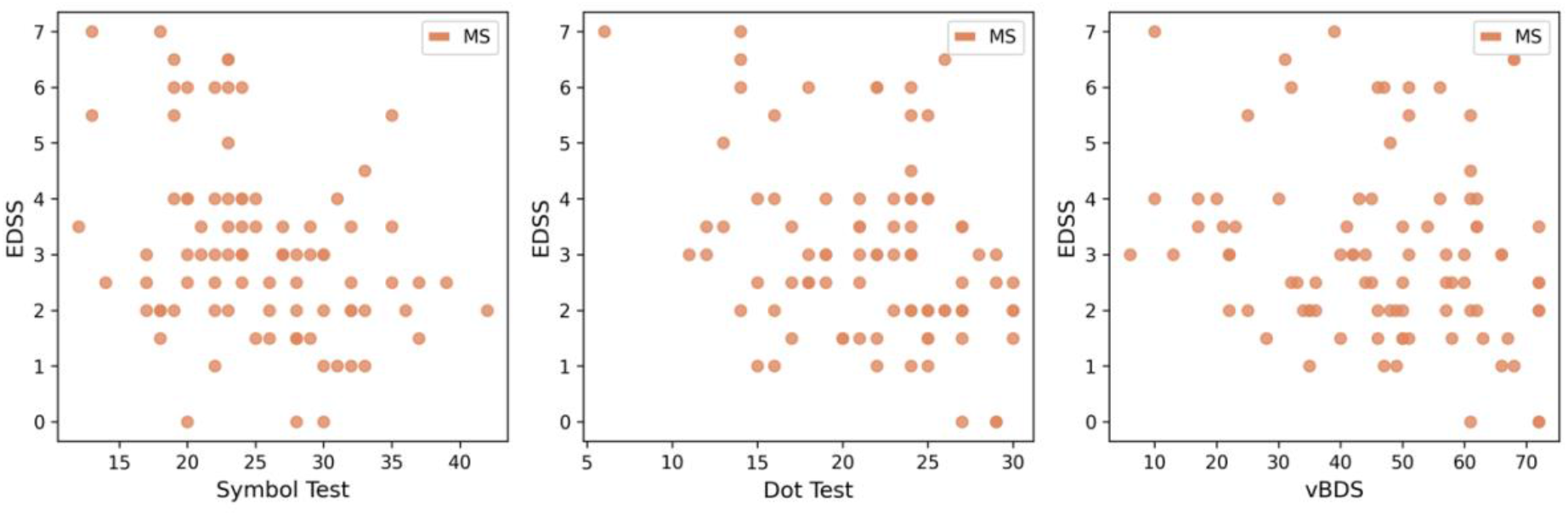
Ecological validity with the expanded disability status scale (EDSS).

## Discussion

In this paper, we present the results of the validation process of a smartphone-based screening battery for cognitive problems in persons with MS. All tests correlated with their paper-pencil equivalent (concurrent validity) and disability build-up (ecological validity), and showed robust test-retest reliability. This indicates the suitability of the battery to be used as a screening tool for routine remote follow-up of cognitive problems, which many persons with MS will develop over time.

Nonetheless, results relating to the criterion validity of ico**gnition** were surprising. Persons with MS scored equally well on all tests compared to healthy subjects. However, as can be observed in table 1 and supplementary figure S2, this was also the case for the paper-pencil tests. Indeed, we appear to have included an MS sample with relatively spared cognition. We tested this in a post-hoc analysis using the analysis of variance (ANOVA) method described in Kallner et al. 2017^32^, comparing SDMT performance of our MS sample (mean = 58.5, standard deviation (SD) = 10.0, N = 101 (cfr. table 1)) with previous literature. López-Góngora et al. 2015 report an average SDMT performance of 54.3 (SD = 13.4, N = 237)^33^, while Sousa et al. 2021 mention an average SDMT performance of 53.51 (SD = 11.76, N = 115). We found that subjects with MS included in this study performed significantly better (comparison with López-Góngora et al. 2015: F = 8.01, p = .005; comparison with Sousa et al. 2021: F = 11.1, p = 0.001)^34^.

In the design of ico**gnition**, careful consideration was given to the potential biasing influence of fine motor impairment in MS^18^. Although we also aim to capture this with a smartphone-based test^35^, motor interference was minimised by using two large buttons for the Symbol Test (contrasting digital SDMT variants where a subject has to choose from 9 smaller buttons^14^), and not putting any restrictions on the response time in the other ico**gnition** tests. Furthermore, ico**gnition** is a quick screening battery with a total testing time of about 15 minutes, similar to the popular Brief International Cognitive Assessment for MS (BICAMS)^16^. Moreover, it reduces clinical workload compared to the classical paper-pencil tests and can be performed whenever the patient feels ready for it (e.g. in terms of fatigue).

Although telemedicine is attractive from a time-management, financial and ecological point of view, it introduces new challenges to which ico**gnition** is no exception. Challenges for the use in a real-world context are confounding factors that can be controlled in a study environment, such as the type of smartphone (e.g. screen size), handling of the smartphone, noise and other distracting circumstances. This can be partly addressed in the application by asking the subject to comply with certain criteria, such as the absence of the aforementioned distractors. Another option could be to take the average performance after repeated testing in a short time-span, in which cognition is not likely to already have deteriorated. Nonetheless, a follow-up study is still warranted to externally validate ico**gnition** in real-world circumstances and using different smartphones. In such a study, careful consideration should also be given to the biasing influence of people’s ability to interact with a smartphone.

We were unable to calculate the traditional WAIS-IV digit span score, as we chose a different design with a fixed number of digit spans to counteract issues in traditional testing^24^. In the aBDS, we used spans from length 3 to 7 to cover a wide range of span lengths. However, as ico**gnition** was designed for people with MS, we redistributed the total number of trials to better match the average backwards digit span score of people with MS, which were previously found to be 4 (Andrade et al. 1999^36^) and 4.67 (Duman et al. 2022^37^). For the scoring of vBDS and aBDS, we also ran a post-hoc test using the total number of correct digits as outcome metric, which did not alter the interpretation of digit span results. Furthermore, in the initial version of the Dot Test, the final 5 trials included remembering the position of the three dots on a 5×5 grid. After testing 3 subjects with MS, we however decided to consistently use a 4×4 grid given the difficulty of the task. Including these 3 subjects in a post-hoc analysis did, however, yield similar results.

## Conclusion

This study presents a newly developed smartphone app, ico**gnition**. It was found to be a valid and reliable tool for remote screening of cognitive functioning in persons with MS. This allows for regular follow-up of cognitive performance to more quickly respond to deterioration.

## Availability of data and code

The anonymised source data on which this paper relies, as well as the code used for statistical analysis and the creation of figures, will be made available in the GitHub repository of our lab: https://github.com/AIMS-VUB/smartphone_tests.

## Data Availability

The data upon which this study relies will be made publicly available in the GitHub repository of our lab: https://github.com/AIMS-VUB/smartphone_tests

## Acknowledgements

We would like to thank Steve De Backer for his efforts in the development of the ico**gnition** app, as well as Eva Keytsman and Florine Wöhler for their support in recruiting and testing subjects for this study.

