## supplementary material for "icognition: a smartphone-based cognitive screening battery"

### Z-normalisation

#### Procedure

To normalise the ico**gnition** test scores for age, sex (male: 1, female: 2) and education level (years of education), for each ico**gnition** test, we fitted a linear regression equation with the test performance as dependent variable and age, sex and education level as independent variables. We then predicted the expected test score of HC and MS subjects given the healthy control regression equation, and subtracted the predicted value from the true test score, yielding the prediction error ($\varepsilon_{pred}$).

$$test\_score_{pred} = w_0 + w_1 * age + w_2 * sex + w_3 * education\_level \qquad \text{(eq 1)}$$

$$\varepsilon_{pred} = test\_score_{norm} - test\_score_{pred} \qquad \text{(eq 2)}$$

For each subject, the z score is the prediction error normalised with respect to the standard deviation of the prediction error distribution of the healthy control subjects.

$$z = \frac{\varepsilon_{pred}}{std(\varepsilon_{pred,HC})} \qquad \text{(eq 3)}$$

#### Necessary values

The necessary information to perform the procedure above is included below.

| | *Symbol Test* | *Dot Test* | *Backwards Digit Span* |
|---|---|---|---|
| $w_0$ | 29.8987 | 28.8806 | 17.0996 |
| $w_1$ | 0.2170 | -1.5233 | 1.2987 |
| $w_2$ | 0.5795 | 0.1559 | 2.5647 |
| $w_3$ | -0.2849 | -0.1610 | -0.2841 |
| $std(\varepsilon_{pred,HC})$ | 4.4741 | 4.1522 | 8.3755 |

*Table S1: Values for the normalisation procedure per ico**gnition** test*

#### Criterion validity on normalized test scores

Figure 1 displays the criterion validity of each ico**gnition** test after correcting test performance for expected performance from the healthy control dataset. People with MS and healthy controls (HC) also scored equally well on these variables (Symbol Test (Z): U = 4367, p = 0.192; Dot Test (Z): U = 3780, p = 0.683; vBDS (Z): U = 3623, p = 0.386).

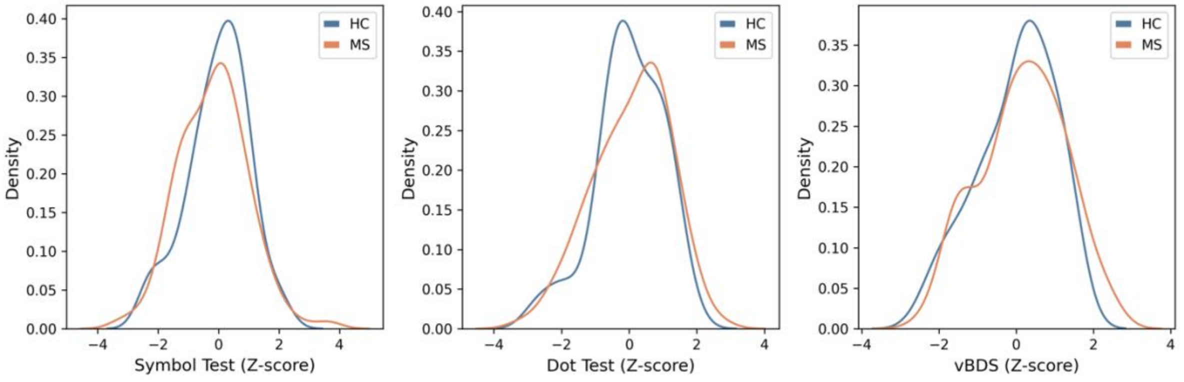


*Figure S1: Criterion validity on normalized test scores*

### Criterion validity paper-pencil tests

None of the paper-pencil tests were significantly different between people with MS and HC subjects (SDMT: U = 3794.5, p = 0.715; SPART 10/36: U = 3792, p = 0.709; auditory Digit Span Backwards (aBDS): U = 3568.5, p = 0.304). The criterion validity (MS versus HC) for all paper-pencil test equivalents of all ico**gnition** tests can be consulted in figure S2.

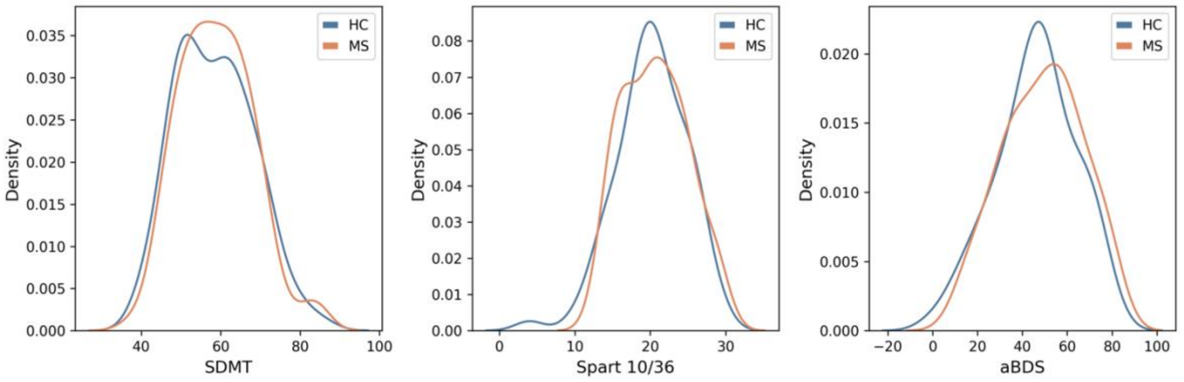


*Figure S2: Criterion validity of paper-pencil tests.*

#### Ecological Validity

This section displays the ecological validity of each ico**gnition** test with disease duration, the Beck Depression Index (BDI) and the Fatigue Scale for Motor and Cognitive functions (FSMC). The Symbol Test did not correlate with any of these variables. The Dot Test was significantly correlated to disease duration (r = -.28, p = .006) and the FSMC total score (r = -.26, p = .012), while the visual Backwards Digit Span was significantly correlated to disease duration (r = -.31, p = .002).

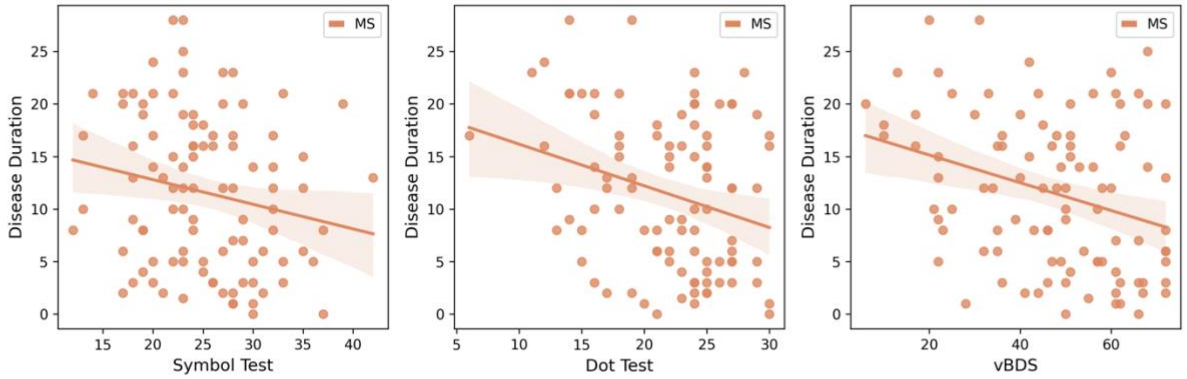


*Figure S3: Ecological validity with disease duration (years)*

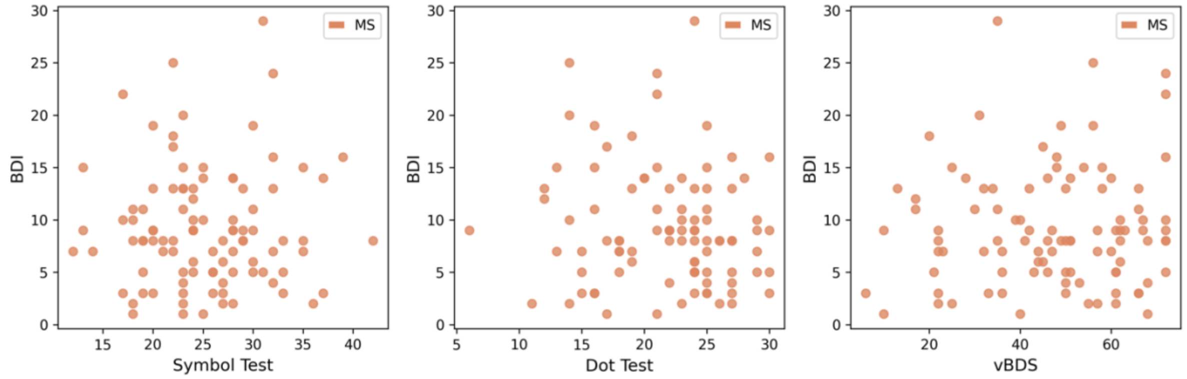


*Figure S4: Ecological validity with the Beck Depression Index (BDI)*

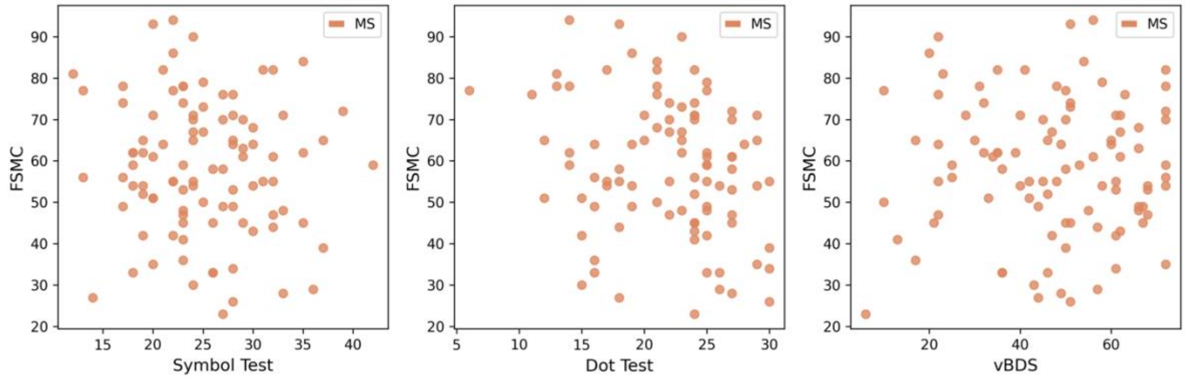


*Figure S5: Ecological validity with the Fatigue Scale for Motor and Cognitive functions (FSMC)*

#### Digit spans

The digit spans used in the auditory Backwards Digit Span are listed in table S1.

| Span length | Digit span |
|---|---|
| 3 | 927 |
| 3 | 843 |
| 4 | 3164 |
| 4 | 5360 |
| 4 | 7918 |
| 4 | 4895 |
| 5 | 28452 |
| 5 | 26019 |
| 5 | 30475 |
| 5 | 84857 |
| 6 | 213604 |
| 6 | 918536 |
| 6 | 639271 |
| 6 | 728938 |
| 7 | 2395480 |
| 7 | 5837686 |

*Table S1: Digit spans used in the auditory backwards digit span*
